# Multi-omics provide evidence for an anti-inflammatory immune signature and metabolic alterations in patients with Long COVID Syndrome – an exploratory study

**DOI:** 10.1101/2022.07.11.22277499

**Authors:** Johannes J. Kovarik, Andrea Bileck, Gerhard Hagn, Samuel M. Meier-Menches, Tobias Frey, Anna Kaempf, Marlene Hollenstein, Tarik Shoumariyeh, Lukas Skos, Birgit Reiter, Marlene C. Gerner, Andreas Spannbauer, Ena Hasimbegovic, Doreen Schmidl, Gerhard Garhöfer, Mariann Gyöngyösi, Klaus G. Schmetterer, Christopher Gerner

## Abstract

Despite the increasing prevalence of patients with Long Covid Syndrome (LCS), to date the pathophysiology of the disease is still unclear, and therefore diagnosis and therapy are a complex effort without any standardization. To address these issues, we performed a broad exploratory screening study applying state-of-the-art post-genomic profiling methods to blood plasma derived from three groups: 1) healthy individuals vaccinated against SARS-CoV-2 without exposure to the full virus, 2) asymptomatic fully recovered patients at least three months after SARS-CoV-2 infection, 3) symptomatic patients at least 3 months after a SARS-CoV-2 infection, here designated as Long Covid Syndrome (LCS) patients. Multiplex cytokine profiling indicated slightly elevated cytokine levels in recovered individuals in contrast to LCS patients, who displayed lowest levels of cytokines. Label-free proteome profiling corroborated an anti-inflammatory status in LCS characterized by low acute phase protein levels and a uniform down-regulation of macrophage-derived secreted proteins, a pattern also characteristic for chronic fatigue syndrome (CFS). Along those lines, eicosanoid and docosanoid analysis revealed high levels of omega-3 fatty acids and a prevalence of anti-inflammatory oxylipins in LCS patients compared to the other study groups. Targeted metabolic profiling indicated low amino acid and triglyceride levels and deregulated acylcarnithines, characteristic for CFS and indicating mitochondrial stress in LCS patients. The anti-inflammatory osmolytes taurine and hypaphorine were significantly up-regulated in LCS patients. In summary, here we present evidence for a specific anti-inflammatory and highly characteristic metabolic signature in LCS which could serve for future diagnostic purposes and help to establish rational therapeutic interventions in these patients.

**One sentence Summary:** Multi-omics plasma analyses demonstrate anti-inflammatory and hypo-metabolic signatures in patients with Long COVID Syndrome.

## Introduction

The outbreak of the COVID-19 pandemic in late 2019 has led to an unprecedented worldwide health crisis with rapid spread of a novel pathogenic member of the coronavirus family (termed SARS-CoV-2) infecting more than 500 million people worldwide. Acute SARS-CoV-2 infection may induce an inappropriate and unique inflammatory response causing the pathognomonic severe respiratory symptoms which can be further accompanied by damage of multiple organs such as brain, heart and kidneys (*1-3*). Accordingly, acute COVID-19 infection may cause high lethality claiming more than 6 million deaths worldwide so far (www.who.int/emergencies/diseases/). Since the start of the pandemic, it has also become evident, that not all patients fully recover following SARS-CoV-2 infection. At first, the observed symptoms were mainly attributed to psychological conditions such as anxiety and stress in the affected individuals (*4*). However, it is now recognized that chronic persistence of COVID-19 symptoms after acute infection constitutes a novel somatic disease entity termed post-acute COVID syndrome (PACS) or Long-COVID Syndrome (LCS) (*5*). Typically, LCS patients suffer from general fatigue, lack of concentration (self-described as “brain fog”) and physical fitness, dyspnea, postural tachycardia as well as a broad range of other clinical symptoms throughout the whole organism, which severely impedes the quality of life (*6*). These clinical presentations mirror the situations found in chronic fatigue syndrome (CFS) which can be secondary to infection with different viruses and were also found in other past coronavirus epidemics including Middle East Respiratory Syndrome (MERS) and Severe Acute Respiratory Syndrome (SARS). Especially for the latter symptom persistence for up to two years has been observed (*7*). Strikingly, development of LCS is not associated with disease severity and so far, only a few studies about potential risk factors and associated comorbidities have been published (*8, 9*). Similarly, the pathogenesis and pathophysiology of this disease remains rather elusive to date. The limited set of available studies on LCS have indicated tissue damage accompanied by chronic inflammation following recovery from acute COVID-19 infection exacerbation of pre-existing (auto)immune-pathologies or persistence of SARS-CoV-2 at distinct sites in the body (*10*). Yet, most of these studies remain rather associative and neither general consent about the predisposition leading to LCS nor consent about the optimal therapeutic management of the disease has emerged so far. This may also be owed to the rather heterogeneous presentation of symptoms and the lack of awareness for this condition, which allows the speculation that most cases so far remain unrecognized. In the light of the high infection rate worldwide, it can be expected that the prevalence of LCS will massively increase in the future, leading to a further COVID-19 associated long-term challenge and burden for the healthcare system. Accordingly, a definition of biomarkers for the diagnosis of LCS as well as in depth studies for the characterization of pathophysiological processes are clearly warranted. Thus, we here set up an exploratory study to perform broad scale mass spectrometry-based multi-omics analysis of blood specimen from healthy donors after vaccination (termed healthy; H), COVID-19 patients who had fully recovered (termed recovered; R) and compared them to samples of LCS patients (termed long-covid; LC) The combination of proteomics and metabolomics focusing on oxylipin analyses has already been demonstrated to strongly support the investigation of pathomechanisms in various diseases (*11-13*). Using this versatile analysis approach, we were able to identify anti-inflammatory and hypo-metabolic signatures in the proteome, lipidome and metabolome of LCS patients, thereby providing insights into the molecular regulation and pathophysiology of LCS.

## Results

### Patient characteristics and study design

The first study cohort consisting of healthy individuals with no history of SARS-CoV-2 infection was recruited three months after full anti-SARS-CoV-2 immunization which was also confirmed by anti-N (-)/anti-S (+) status (termed healthy; H). The second recruited cohort consisted of age- and gender-matched individuals who had a SARS-CoV-2 infection history at least three months prior to inclusion into this study, but were symptom free in anamnesis at the time of blood draw (termed recovered; R). Infection status was confirmed by positive anti-N and anti-S antibody levels. The third group of study patients had similarly succumbed to PCR positive SARS-CoV-2 infection at least three months before presentation. All patients in this group had a positive anamnesis of chronic fatigue and/or severe chronic lack of concentrations following SARS-CoV-2 infection combined with at least one more chronic symptom including dyspnea, coughing and loss of smelling among others at the time of presentation, qualifying them for the diagnosis LCS (whole patient characteristics are displayed in Table 1A and 1B). Again, post infection status was confirmed by positive antibody testing for anti-N and anti-S antibodies. For the here described exploratory study, specimen from 13 individuals from each group were selected for broad scale multi-omics analyses including a cytokine array, untargeted shotgun proteomics, an untargeted eicosanoid/docosanoid analysis and a comprehensive commercial metabolomics kit. Routine laboratory testing of basic protein, lipid and lipoprotein parameters showed no differences between the three groups. Similarly, CRP levels in all three groups were below or only minimally above the threshold (Table 2). Furthermore, none of the analyzed individuals presented with fever or displayed any clinical signs of systemic infection at the time of blood withdrawal, ruling out major systemic metabolic or acute inflammatory processes in the tested individuals at this time point.

**Table 1:** Study group characteristics.

| **Table 1A** | **healthy (n=13)** | **recovered (n=13)** | **long-covid (n=13)** |
| --- | --- | --- | --- |
| **General data** |  |  |  |
| Gender (f:m) | 7:6 | 7:6 | 9:4 |
| Age [years] | 30 (25-43) | 32 (24-49) | 33 (21-53) |
| Time after exposure [months] | 6 (3-8) | 10 (3-12) | 7 (3-10) |
| **Chronic disease** (total number (percentage)) |  |  |  |
| Asthma bronchiale | 0 | 1 (0.07) | 4 (0.31) |
| Multiple sclerosis | 0 | 0 | 1 (0.07) |
| Autoimmune thyroiditis | 0 | 1 (0.07) | 1 (0.07) |
| Atopic dermatitis | 0 | 1 (0.07) | 0 |
| Psoriasis arthritis | 1 (0.07) | 0 | 0 |
| **Table 1B** | **healthy (n=13)** | **recovered (n=13)** | **long-covid (n=13)** |
| **Chronic Symptom** (total number (percentage)) |  |  |  |
| Lack of concentration | - | - | 10 (0.77) |
| Fatigue | - | - | 9 (0.69) |
| Dyspnoea | - | - | 9 (0.69) |
| Chronic cough | - | - | 2 (0.15) |
| Muscular aching | - | - | 3 (0.23) |
| Amnesic aphasia | - | - | 5 (0.38) |
| Sleep disorder | - | - | 4 (0.31) |
| Urinary incontinence | - | - | 2 (0.15) |
| Nausea | - | - | 3 (0.23) |
| Tinnitus | - | - | 4 (0.30) |
| Other symptoms | - | - | 9 (0.69) |

**Table 2:**
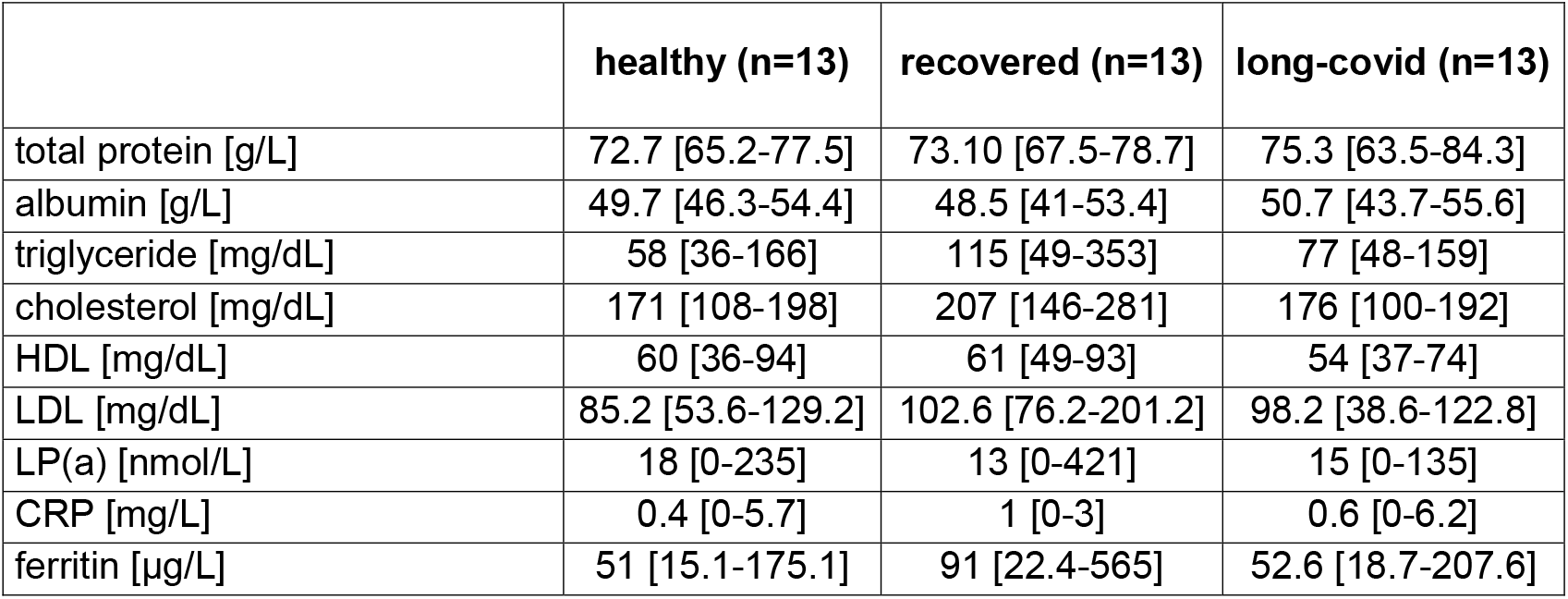
Routine serum parameters.

|  | **healthy (n=13)** | **recovered (n=13)** | **long-covid (n=13)** |
| --- | --- | --- | --- |
| total protein [g/L] | 72.7 [65.2-77.5] | 73.10 [67.5-78.7] | 75.3 [63.5-84.3] |
| albumin [g/L] | 49.7 [46.3-54.4] | 48.5 [41-53.4] | 50.7 [43.7-55.6] |
| triglyceride [mg/dL] | 58 [36-166] | 115 [49-353] | 77 [48-159] |
| cholesterol [mg/dL] | 171 [108-198] | 207 [146-281] | 176 [100-192] |
| HDL [mg/dL] | 60 [36-94] | 61 [49-93] | 54 [37-74] |
| LDL [mg/dL] | 85.2 [53.6-129.2] | 102.6 [76.2-201.2] | 98.2 [38.6-122.8] |
| LP(a) [nmol/L] | 18 [0-235] | 13 [0-421] | 15 [0-135] |
| CRP [mg/L] | 0.4 [0-5.7] | 1 [0-3] | 0.6 [0-6.2] |
| ferritin [µg/L] | 51 [15.1-175.1] | 91 [22.4-565] | 52.6 [18.7-207.6] |

To confirm our hypothesis regarding the role of fatty acids in LCS (see below) and to exclude an effect from oral supplementation, we analyzed samples from a fourth cohort including 10 healthy subjects which had not been exposed to SARS-CoV-2. For this purpose, these subjects received tablets containing 870mg Omega-3 (420 mg EPA and 330 mg DHA) twice daily in a prospective study design for one week and plasma samples were obtained before start of intake and after intake of the last dose.

### Immune activation marker profiling displays a lack of systemic inflammation in LCS patients

In order to investigate whether LCS may result from still unresolved inflammatory processes after the viral infection, a broad panel of 65 cytokines, chemokines and soluble receptors associated with immune activation was assayed. Remarkably, neither pro-nor anti-inflammatory cytokines were found significantly up-regulated in LCS patients compared to the other two groups. However, striking was the apparent down-regulation of IL-18 in LCS patients, as this T-cell and macrophage derived pro-inflammatory cytokine is orchestrating migration and antiviral response of macrophages (Figure 1A) Furthermore, the monocyte/macrophage-derived factors MCP-1/CCL-2 and sTNF-RII, were found significantly down-regulated in LCS patients compared to fully recovered patients (Figure 1A). Overall, cytokine levels were rather low in LCS patients, indicating a lack of pro-inflammatory activities.

**Figure 1:**
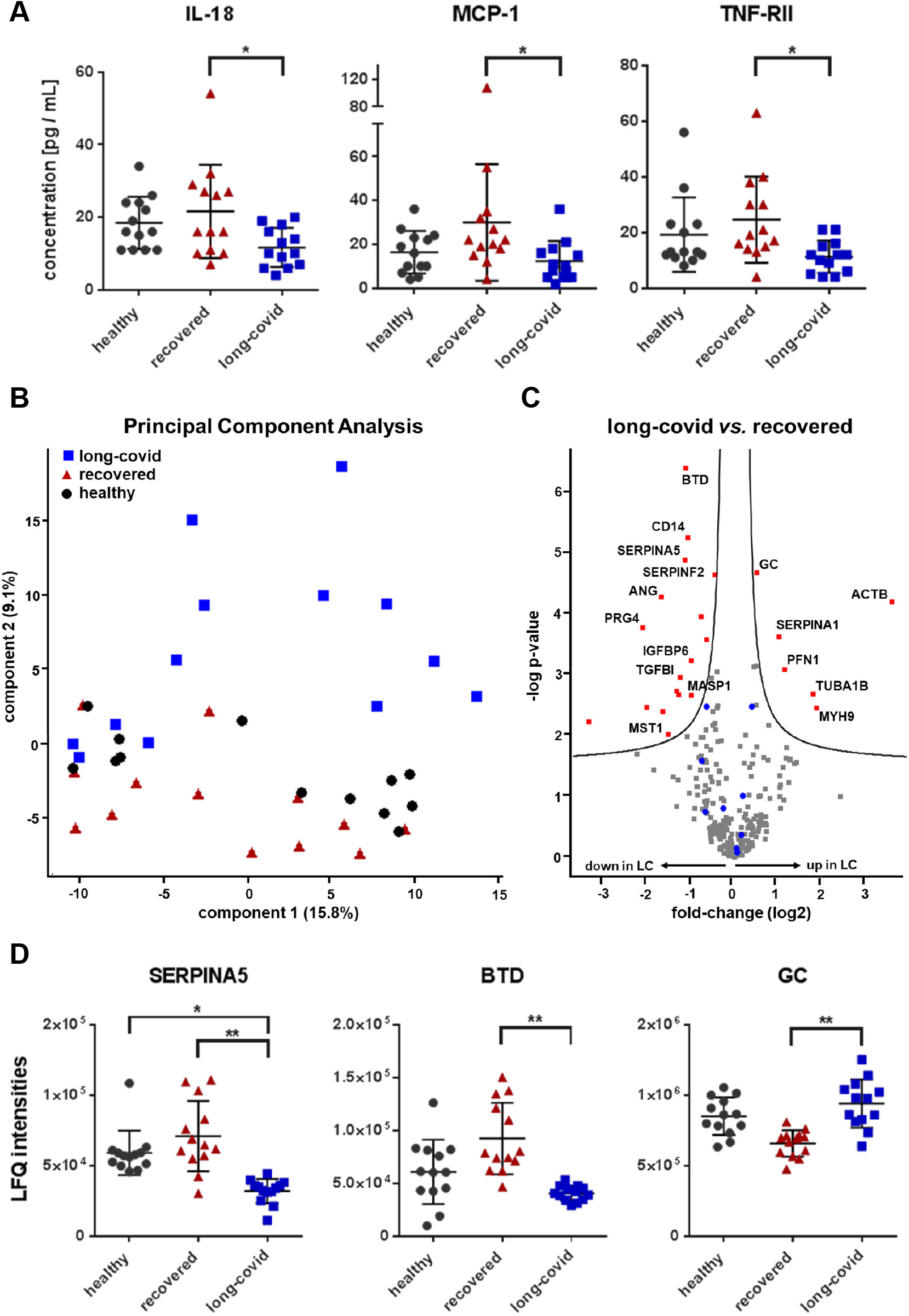
Cytokine and proteome profiles demonstrate a lack of pro-inflammatory activities in long-covid patients. **(A)** Significant differences of IL-18 as well as the macrophage-derived cytokines MCP-1 and TNF-RII between long-covid and fully recovered patients are depicted. **(B)** A Principal Component Analysis based on proteome profiling of plasma samples from long-covid patients (blue square), fully recovered patients (red triangle) and vaccinated healthy controls (grey circle) is shown. **(C)** Significant differences in plasma protein abundance between long-covid and fully recovered patients are visualized by a volcano plot. **(D)** Label-free quantification (LFQ) intensities derived from untargeted plasma proteomics of SERPINA5, Biotinidase (BTD) and Vitamin D-binding protein (GC) are depicted for all study groups. * p ≤ 0.05, ** p ≤ 0.01

### Proteome profiling displays and anti-inflammatory pattern in LCS patients

To further elucidate the inflammatory status in patients with LCS, we performed shotgun proteomics of plasma samples of the three study groups. Principle component analysis (PCA) distributed the proteome profiles of healthy vaccinated individuals mainly between the almost completely separated groups of LCS and recovered patients (Figure 1B, Supplementary Figure S1, Supplementary Table S1). Thus, significant (FDR < 0.05) proteome alterations were mainly observed comparing the latter two groups (Figure 1C). Apart from several cell leakage products typically derived from uncontrolled cell death, including actin (ACT), tubulin (TUBA1B), myosin-9 (MYH9) and profilin (PFN1), only alpha-1-anti trypsin (SERPINA1) and vitamin D binding protein (GC; Figure 1D) were found significantly up-regulated in the LCS group. A larger number of proteins was found down-regulated, including the protease inhibitors SEPINA5 and SERPINF2, Biotinidase (BTD; Figure 1D) and the macrophage-associated proteins soluble CD14, ANG and Proteoglycan 4 (PRG4). The acute phase protein CRP was only detected (near threshold levels) in a few samples and thus removed upon filtering. Other acute phase proteins including serum amyloid A and serum amyloid P component, fibrinogens, orosomucoid and alpha-2-macroglobulin were readily detectable but rather down-regulated in LCS patients. These findings further demonstrate the absence of systemic inflammation in LCS patients and potentially indicate shifts in the activation of inflammatory immune subsets.

### Fatty acid and oxylipin analysis indicated increased phospholipase A2 activities affecting preferentially mitochondrial lipids

Oxidized products of polyunsaturated fatty acids such as arachidonic acid (AA) represent highly bioactive but short-lived signaling molecules and lipid mediators, often termed eicosanoids. Phospholipase A2 catalyzes the first step of biosynthesis, the release of polyunsaturated fatty acids from cell membrane phospholipids. The bioactive oxylipins are subsequently formed by the action of cyclooxygenases, lipoxygenases or cytochrome P450 monooxygenases and modulate numerous physiological functions including bronchial and vascular tonus, thrombocyte function, inflammation and other immune responses (*14*). Thus, assessment of the lipidome allows insights into diverse physiological and pathophysiological processes. Following the analysis of plasma eicosanoids from the three study groups, PCA of fatty acids and their oxidation products rather separated healthy controls from LCS patients, with the group of recovered patients dispersed in between (Figure 2A, Supplementary Table S2). Thus, here most of the significant events were observed comparing LCS patients to healthy controls (Figure 2B). Evidently, higher levels of all kinds of polyunsaturated fatty acids were characteristic for LCS, pointing to higher phospholipase A2 activities. However, increased plasma levels of the rather pro-inflammatory AA were mainly observed in the recovered group (Figure 2C). In contrast, LCS was marked by the predominant release of the anti-inflammatory molecules EPA and DHA into the blood of affected patients (Figure 2B, C). Plasma levels of these molecules may derive from intracellular sources but can also be affected by confounders such as nutrition. Two independent observations indicate that this was not the case in the studied LCS-cohort. First, the DHA oxidation products 17- and 22-HDoHE were also found significantly increased in LCS patients (Figure 2D). Second, polyunsaturated fatty acids may occur together with their trans-isoforms (*15*). In an independent cohort of 10 healthy volunteers, we observed that nutritional supplementation of DHA was not associated with increased levels of trans-DHA (Figure 2E). In contrast, the levels of trans-DHA were consistently increased in LCS patients, suggesting that DHA was preferentially released from intracellular sources. Accordingly, the overall ratio of Ω3-fatty acids to Ω6-fatty acids was found significantly increased in LCS patients (Figure 2F). A general anti-inflammatory lipid mediator pattern in LCS patients was also corroborated by increased levels of the 15-LOX product from linoleic acid, 13-OxoODE (*16*), when compared to recovered patients (Figure 2E). Thus, here we provide evidence that increased plasma levels of anti-inflammatory oxylipins may be a characteristic feature for LCS patients.

**Figure 2:**
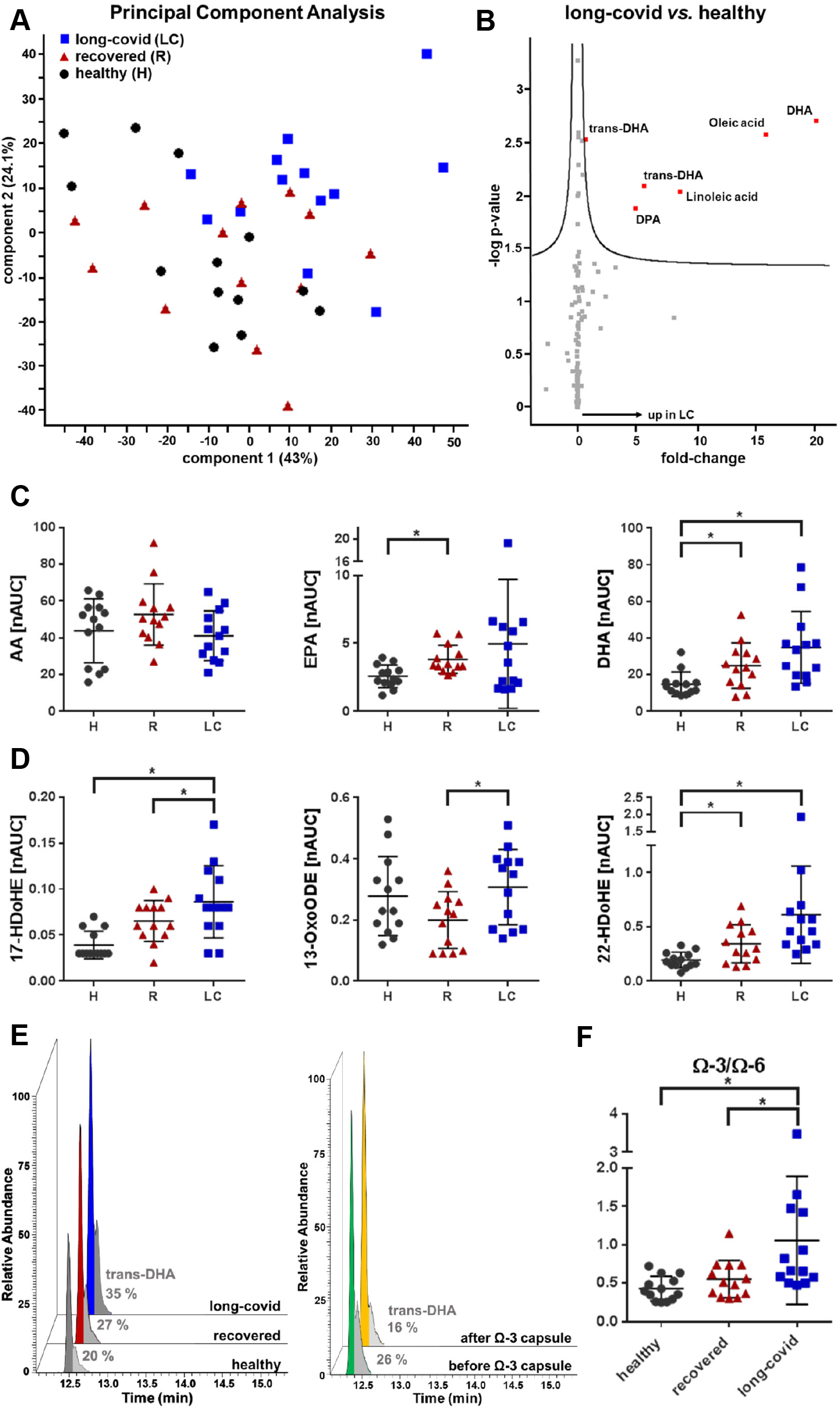
Increased levels of fatty acids with a prevalence for Ω3-fatty acids and anti-inflammatory docosanoids is characteristic for long-covid. **(A)** A Principal Component Analysis based on eicosanoid analysis of plasma samples from long-covid patients (blue square), fully recovered patients (red triangle) and vaccinated healthy controls (grey circle) is shown. **(B)** Significant differences in plasma eicosanoids between long-covid patients and vaccinated healthy controls are visualized by a volcano plot. **(C, D)** Normalized area under the curve (nAUC) values of arachidonic acid (AA), eicosapentaenoic acid (EPA), docosahexaenoic acid (DHA) and the eicosanoids 17-HDoHE, 13-OxoODE as well as 22-HDoHE are depicted for each study group (H, vaccinated healthy controls; R, fully recovered patients; LC, long-covid patients). **(E)** Increased levels of trans-DHA in long-covid patients are shown in an extracted ion chromatogram (m/z = 327.233 at a retention time (Time) of 12.5 min). No increase in trans-DHA levels after nutritional supplementation of omega-3 (Ω-3) capsules was observed in healthy controls. Percentage (%) of trans-DHA to DHA is depicted in grey for each study group. **(F)** A significant increase in the omega-3 to omega-6 ratio (Ω-3/ Ω-6) was observed in long-covid patients compared to vaccinated healthy controls as well as fully recovered patients. * p ≤ 0.05

### Metabolomic aberrations relate to chronic fatigue syndrome and display an osmolyte-mediated anti-inflammatory signature

In case of metabolites, PCA analysis indicated maximal contrast between recovered and LCS patients (Figure 3A) similar to the plasma protein analyses described above. Indeed, out of 474 metabolites and lipids analyzed, 107 were found significantly deregulated between these two groups (Figure 3B, Supplementary Table S3). Most apparently, a large number of metabolic alterations were related to a disturbance in energy metabolism, as evidenced by the down-regulation in the LCS group of branched chain amino acids Val, Leu and Ile, the aromatic amino acid Tyr and amino acids Trp and Arg which also act as inflammation mediators (Figure 3C, Supplementary Figure S2). Furthermore, increased levels of C18:1-acylcarnitine accompanied with a down-regulation of C3-acylcarnitine were observed in LCS patients (Figure 3C). This pattern may indicate reduced levels of beta oxidation accompanied by increased amino acid catabolism. A further indication for a disturbed energy metabolism was the significant increase in lactate, pointing to increased anaerobic glycolysis (Supplementary Figure S2, Supplementary Table S3). Furthermore, the majority of lipids including various triacylglycerols, glycosylceramides, glycerophospholipids and several ceramides were found down-regulated in LCS (Figure 3D, Supplementary Table S4), which bears resemblance to a pattern characteristic for chronic fatigue syndrome (*17*). Remarkably, an LCS-associated lack of inflammatory processes was also evidenced by the present metabolomics results. The metabolite most significantly up-regulated in LCS patients was hypaphorine (TrpBetaine), an anti-inflammatory alkaloid (*18*) described to induce sleep in mice (*19*). Hypaphorine may act as osmolyte similar to the anti-inflammatory molecule taurine (Figure 3C), which was also significantly up-regulated in LCS. In contrast, hypoxanthine levels were strongly increased in the recovered group while LCS patients showed comparable levels to the healthy vaccinated group.

**Figure 3:**
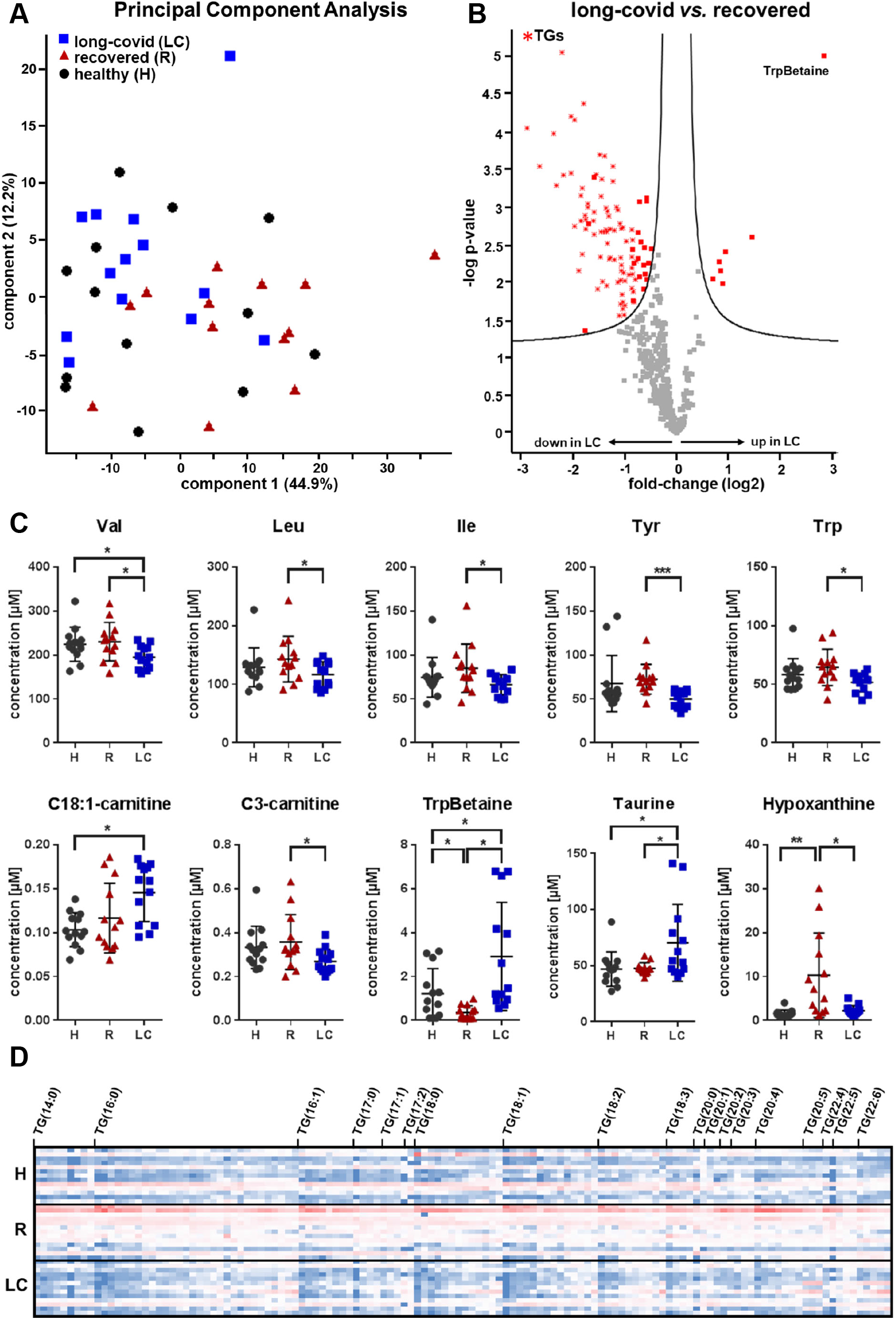
Plasma metabolomics reveals disturbances in the energy metabolism of long-covid patients. **(A)** A Principal Component Analysis based on metabolomic analysis of plasma samples from long-covid patients (blue square), fully recovered patients (red triangle) and vaccinated healthy controls (grey circle) is shown. **(B)** Significant higher levels of TrpBetaine as well as significant lower levels of triacylglycerols (TGs, red star) in plasma samples of long-covid patients are visualized by a volcano plot. **(C)** Plasma levels of the amino acids tyrosine (Tyr), tryptophan (Trp), valine (Val), leucine (Leu) and isoleucine (Ile) as well as of carnitines, hypoxanthine, TrpBetaine and taurine are depicted for each study group (H, vaccinated healthy controls; R, fully recovered patients; LC, long-covid patients). **(D)** A heat map of all identified triacylglycerols (TGs) displays higher levels of TGs in fully recovered patients compared to long-covid patients and vaccinated healthy controls. * p ≤ 0.05, ** p ≤ 0.01, *** p ≤ 0.001

#### Alternatively polarized macrophage-like cells display features of the LCS-associated anti-inflammatory signature

A systemic inhibition of inflammatory processes as presently observed in the LCS cohort may actually result from a predominance of alternatively polarized macrophages (*20*). To address this issue, we investigated a U937 cell-based macrophage *in vitro* model system again applying proteome and eicosanoid/docosanoid analysis. U937 cells were differentiated into macrophages using PMA and polarized into either M1-like cells by LPS or M2-like cells with GMCF and IL-4 (Figure 4A). As expected, the M1-like cells were found to robustly produce pro-inflammatory cytokines such as IL-1beta, CCL1, CXCL2, CXCL5 and other inflammation marker such as MMP9 (Figure 4B). Indeed, two of the cytokines down-regulated in LCS, MCP-1 (CCL2) and TNFRSF1B (see Figure 1A), were found mainly secreted by M1-like cells and rather down-regulated in M2-like cells (Figure 4B). M1-like macrophages also secreted the protease furin (Figure 4B). Furin was described to cleave the SARS-CoV-2 spike protein in order to allow SARS-CoV-2 virus particles to enter human cells. The main furin inhibitor, SERPINA5, was found significantly down-regulated in LCS patients (Figure 1D), pointing to a relevant pathomechanism. The M2-like cells displayed a tolerogenic phenotype illustrated by the expression of the immune suppressor CCL18 (*21*) and the angiogenesis promoting factor ANGPT4. In addition, the M2-like cells expressed several proteins promoting lipolysis (PLA2G4A), altered lipid metabolism (PLIN2 and FABP4) and lipid peroxidation (HMOX1) (Figure 4C). HMOX1 is actually both an essential enzyme for iron-dependent lipid peroxidation during ferroptotic cell death (*22*) and an antiviral protein (*23*). Polarized macrophages also released fatty acids and their oxidation products. Both arachidonic acid and DHA were released by M2-like cells more than by M1-like cells (Figure 4D). In line with the specific expression of COX2 (PTGS2) in M1-like cells, the COX-products PGE2, PGF2alpha and TXB2 were only detected in M1-like cells (Figure 4D). On the other hand, the lipid peroxidation products HpODE and the cytochrome P450 product 12,13-DiHOME were found at higher levels released from M2-like cells. Thus, the M2-macrophage pattern observed *in vitro* rather resembles the situation found in LCS patients.

**Figure 4:**
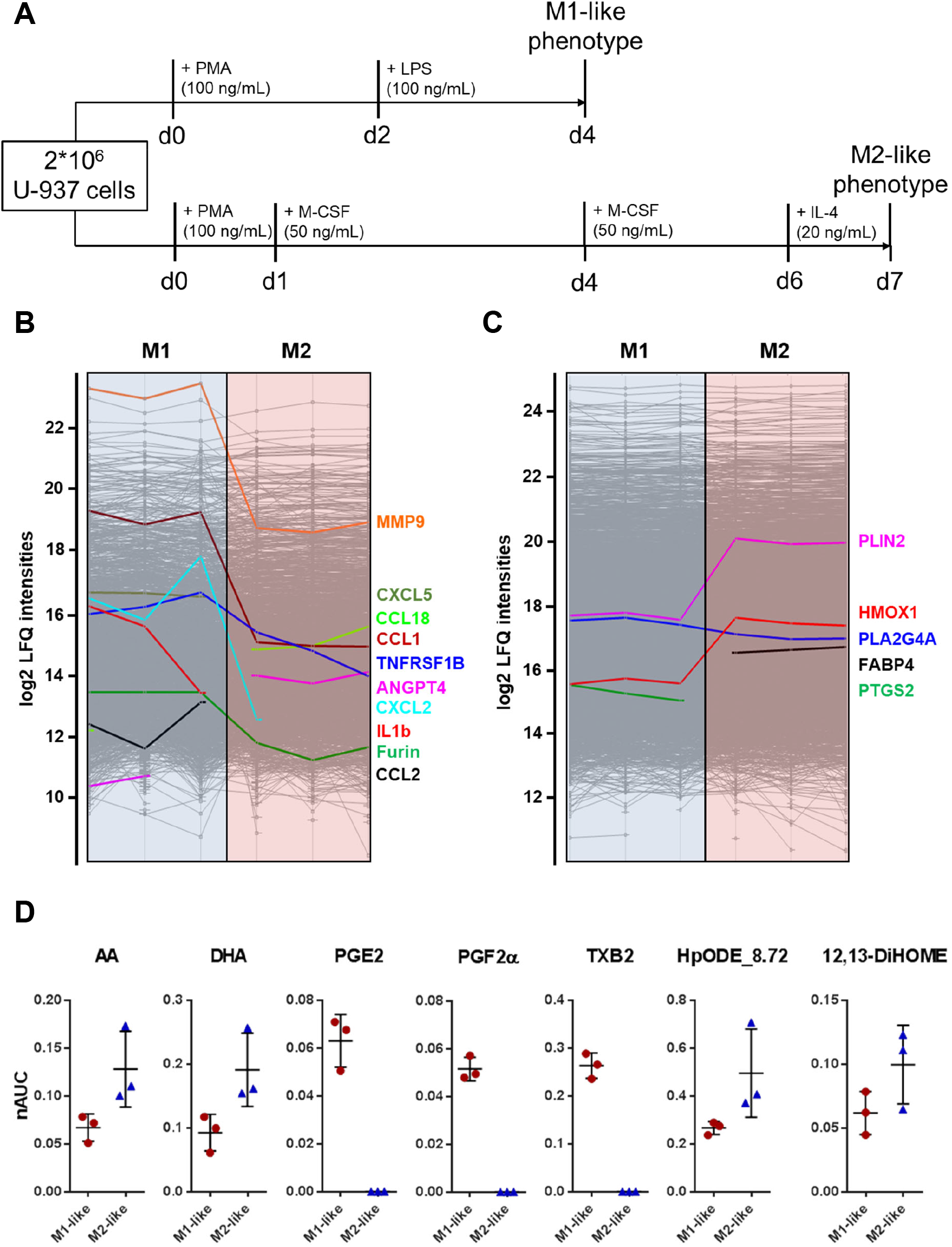
Multi-omcis of in vitro polarized macrophages reveal molecular features similar to the LCS. **(A)** Scheme of *in vitro* polarization of U-937 cells to either M1-like or M2-like macrophages (d, days). Log2-transformed label-free quantification (LFQ) intensities derived from untargeted proteomics of **(B)** the secretome and **(C)** the cell lysates of M1-like macrophages (M1) and M2-like macrophages (M2) are depicted using profile plots. **(D)** Normalized area under the curve (nAUC) values of arachidonic acid (AA), docosahexaenoic acid (DHA) and the eicosanoids PGE2, PGFa, TXB2, HpODE as well as 12,13-DiHOME in the secretome of *in vitro* polarized M1-like and M2-like macrophages are shown.

## Discussion

Post-acute sequelae of SARS-CoV-2 infection (termed Long COVID Syndrome) can be found in about 10% of affected patients and thus pose an ever-increasing burden. So far, the pathophysiology of LCS is unknown and rather the subject to speculative hypotheses such as persisting chronic inflammation after infection (*10*). Given this lack of information we chose to perform a broad scale exploratory study assessing the proteome, lipidome and metabolome in LCS patients. As control groups we recruited individuals who had fully recovered after acute COVID-19 infection and healthy individuals after COVID-19 vaccination. These analyses allowed us to identify LCS specific molecular patterns which strongly point at several unexpected processes.

Numerous studies have clearly established that acute COVID-19 infection is associated with hyperinflammation accompanied by metabolic alterations and increased mitochondrial ROS production. Thus, a failure to clear such inflammatory activities has been commonly assumed to account for LCS symptoms (10). However, in contrast to the acute infection situation, we present conclusive evidence for systemic anti-inflammatory conditions in LCS patients. A lack of pro-inflammatory activities and the predominance of anti-inflammatory mediators in blood plasma was independently observed at the levels of cytokines, acute phase proteins, oxylipins and metabolites. Furthermore, metabolomics analyses also strongly indicated a mainly catabolic metabolism in LCS patients, which may account for the characteristic chronic fatigue symptoms.

Of the detected cytokines, chemokines and soluble receptors, the three markers IL-18, soluble TNF-RII and MCP-1/CCL2 were significantly down-regulated in the LCS group. All three factors have pro-inflammatory functions and reflect the activation and communication of T-lymphocytes and monocytes/macrophages. In the proteome, down-regulation of acute phase proteins was observed, which was most pronounced between the recovered and the LCS group. Of note, SERPINA5 levels were significantly decreased in the LCS cohort compared to both the healthy as well as the recovered group, thus identifying a unique feature of LCS. SERPINA5 serves as antagonist of the protease furin, which is essential for viral entry into human cells (*24*). Thus, it is intriguing to speculate that alteration in the furin/SERPINA5 interplay – either due to genetic background of the individual or due to specific immune responses during SARS-CoV-2 infection – might provide a basis for the development of LCS.

Furthermore, the observed proteome patterns indicate differential monocyte/macrophage polarization and activity between the LCS and the recovered group since the most significantly down-regulated proteins in LCS patients were found to be derived from macrophages or to directly affect macrophage function (Figure 5). CD14 is a macrophage-specific membrane protein eventually secreted into plasma upon inflammatory activation (*25*). Down-regulation of CD14 may thus indicate less M1-like macrophage responses in LCS patients. Angiogenin (ANG) is an angiogenic protein also described as anti-bacterial protein secreted by macrophages (*26*). Proteoglycan-4 (PRG4) is mainly expressed by fibroblast-like cells but serves as an important regulator of inflammation (*27*) and has been described to act as an essential regulator of synovial macrophage polarization and inflammatory macrophage joint infiltration (*28*). Biotinidase has been described to be essential for basic macrophage functions. Intriguingly, biotinidase deficiency may account for hypotonia, lethargy, cognitive retardation, and seizures (*29*) thus potentially linking our observations to the symptom complex of LCS patients. Finally, Vitamin D binding protein (GC), actually up-regulated in plasma of LCS patients, may also be involved in macrophage functions, as it is a precursor for the so-called macrophage-activating signal factor (*30*). Furthermore, among the up-regulated proteins in the LCS group, SERPINA1 has been described as characteristic marker of M2-polarized macrophages (*31*), thus additionally giving evidence about alternative macrophage polarization in LCS.

**Figure 5:**
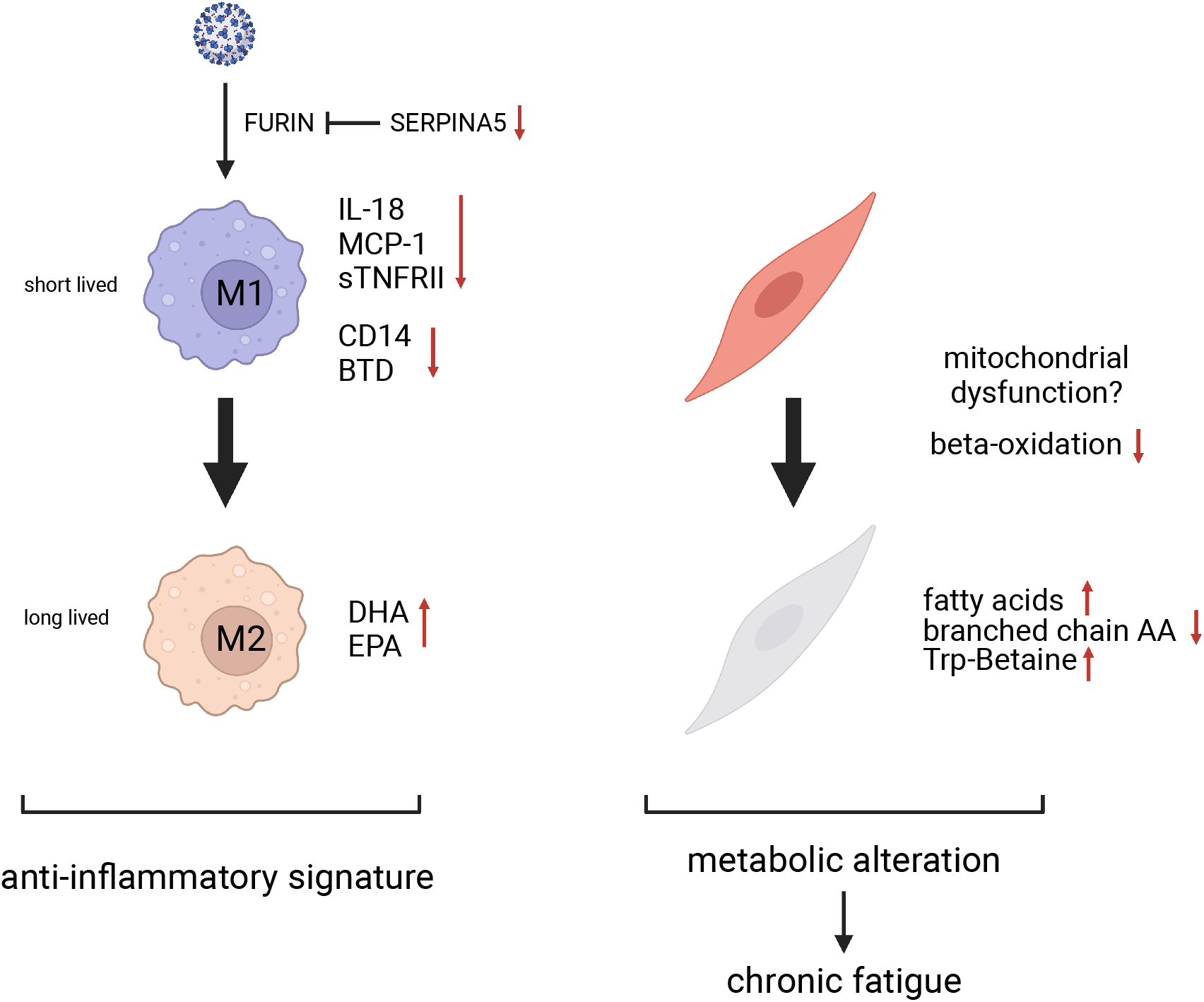
Outline of the suggested pathomechanisms. Monocytes and muscle cells may be specifically affected in LCS patients. In the course of chronic viral infection, there is evidence for a decrease in the occurrence and activities of M1 macrophages accompanied with an increase in M2-like macrophage activities. However, also muscle cells may account for some of the observed alterations and may as well be affected by viral infection *via* mitochondrial dysfunction, indicated by a lack of beta-oxidation accompanied with decreased levels of branched chain amino acids.

Apart from the proteome analyses, also the observed patterns in the lipidome of LCS patients provided independent evidence for an anti-inflammatory status. Along those lines, we identified increased levels of DHA, its metabolites and other docosanoids in LCS patients, which are mainly regarded as anti-inflammatory and may promote tolerogenic macrophage polarization (*32*). Of note, DHA does not only act as anti-inflammatory mediator but high DHA levels are relevant for normal brain function (*33*) as well as mitochondrial functions in different cell types (*34*). DHA and other Ω3-fatty acids are preferentially released by calcium-independent phospholipase iPLA2 (*35*) which is broadly expressed throughout the body and induced upon oxidative stress and mitochondrial damage (*35*). Thus, we hypothesized that the observed increase in Ω3-fatty acids in LCS was a response to oxidative stress generated during the COVID-19 infection. It can again only be speculated whether this is due to a distinct course of SARS-CoV-2 infection in LCS patients or due to individual variations in the anti-oxidative cellular mechanisms. In this respect, longitudinal studies addressing these points in larger cohorts are clearly warranted.

An LCS-associated lack of inflammatory processes was also evidenced by the present metabolomics results. The osmolyte taurine, which has been attributed with anti-inflammatory and anti-oxidative properties (*36*), was significantly increased in LCS patients compared to the other two groups. The metabolite most significantly up-regulated in LCS patients was hypaphorine (TrpBetaine), an anti-inflammatory alkaloid (25) described to induce sleep in mice (26), which might thus also relate to the chronic fatigue symptoms in LCS. Inversely, hypoxanthine levels, which are among others indicative of tissue hypoxia as found during acute inflammation, were strongly increased in the recovered group, while LCS patients showed similar levels to the healthy group.

Apart from this overall pattern of anti-inflammation, the metabolome analyses also provided first indications regarding an aberrant amino acid catabolism in LCS. Along those lines, levels of branched chain amino acids were significantly decreased in the LCS group, including both glucogenic as well as ketogenic amino acids. This observation gives evidence for increased energy consumption from protein breakdown, which is in line with reports of increased muscle weakness and sarcopenia due to metabolic alterations (Figure 5) during SARS-CoV-2 infection (*37*). Furthermore, tryptophan levels were found down-regulated in the LCS group potentially resulting from prolonged IDO activity, which has also been observed during acute SARS-CoV-2 infection (*37*). Metabolic alterations were also found at the level of triacylglycerols and other complex lipids which were rather low in the LCS group. Increased levels of fatty acids accompanied by decreased levels of triacylglcerols point to increased activities of endothelial lipase (23), which was associated with cognitive impairment (24). Intriguingly, this pattern is highly similar to the observations described for CFS (*38*), providing a possible explanation for the fatigue and brain fog symptoms observed in LCS. These findings also underline the potential to introduce novel dietary approaches into tailored rehabilitation regimen. In this regard also strict control of metabolic comorbidities like diabetes, could help in reducing and managing LCS (*39*).

From the above observed molecular patterns an overall contribution of alternatively polarized M2 macrophages may be deduced. During acute infection peripheral blood monocytes are strongly affected (*40*) and are major contributors to the inappropriate inflammatory reaction (*41*). While not contributing to viral replication, macrophages may eventually get infected themselves by SARS-CoV-2 (*42, 43*) and peripheral blood monocytes were described to be significantly altered during COVID-19 infection (*40*). *In vitro* polarized M1-like macrophages express the protease furin (see Figure 4B) which is essential for viral entry into human cells (*24*) and might therefore contribute to the course, severity and long-term sequelae of the infection. However, the pro-inflammatory M1 state may be switched, e.g. by oxidized phospholipids accumulated during acute infection, to a tolerogenic M2-like phenotype (*44, 45*), thus becoming long-lived cells coordinating tissue regeneration (*46*). Lipid peroxidation may occur consequent to inflammatory responses also during sterile inflammation such as atherosclerosis (*47*) and thus represents a plausible mechanism inducing tolerogenic macrophage polarization after SARS-CoV-2 infection. Polarized tolerogenic macrophages suppress pro-inflammatory cytokines (*48*) and induce an anti-inflammatory state (*49*), and are promoted by anti-inflammatory docosanoids (*32*), highly reminiscent to the presently observed molecular pattern characteristic for LCS patients. As a consequence, here we suggest alternative macrophage polarization as key cell mechanism accounting for LCS symptoms. This polarization may also contribute to the observed metabolic alterations since macrophages play a central role for muscle metabolic control as well as regeneration and homeostasis (*50*) (*51*). Along those lines, disturbance of muscle-macrophage crosstalk has been described to accompany obesity (*52*). Thus, aberrant macrophage polarization might also contribute to the metabolic pattern associated with chronic fatigue syndrome (Figure 5). Although there is currently no proof for this potential disease mechanism, this hypothesis provides ample opportunities for rigorous clinical testing. It is fair to expect quite some model refinements consequent to such studies, but already at this stage of research this study points to completely new strategies how to establish highly effective therapies.

The present data also provide insights into the processes of successful recovery after acute SARS-CoV-2 infection. It is noteworthy that the symptom-free recovered individuals showed alterations compared to the healthy control group in many assessed parameters throughout the different biomolecular compartments. Taken together these findings provide evidence that, even months after acute infection, systemic processes are still active in these individuals. Thus, SARS-CoV-2 infection might leave molecular remnants such as infected macrophages long after symptomatic recovery. Three potential mechanisms have been suggested recently to account for LCS: immune dysregulation, autoimmunity or viral persistence (*53*). Our data are fully compatible with immune dysregulation and viral persistence, but hardly support a general role of autoimmunity, as this should be expected to be associated with a pro-inflammatory pattern. While it is evident that the limited cohort size would not allow us to draw general conclusions, it is also evident that the described data are of sufficient power to assess the compatibility with existing hypotheses and to suggest novel ones.

Thus, the obtained molecular patterns do not only provide first insights into the pathophysiology of COVID-19 sequelae as depicted in Figure 5, but may also provide a first basis for the definition of LCS specific biomarkers. Our broad scale analyses could not detect a unique and specific marker for the disease. However, many significant molecular alterations can be clearly associated with characteristic symptoms of the disease. It is possible to hypothesize regarding the biological causes of these alterations. In addition, it can be envisioned that a combination of presently described proteins (e.g. low SERPINA5), docosanoids (e.g. high DHA) and small metabolites (e.g. high hypaphorine) in patients with characteristic anamnesis and symptoms could help to identify and better define LCS. In this regard, large scale studies to assess the potential sensitivity and specificity of such scores, including the consideration of different SARS-CoV-2 strains, are clearly warranted.

In summary, here we present a distinct multi-omics signature demonstrating a prevalence of anti-inflammatory effector molecules combined with molecular patterns of characteristically altered metabolism detectable in plasma of LCS patients, offering a unique chance for diagnosis with selected molecular biomarkers and providing novel hypotheses about the pathophysiology of the disease, thus potentially aiding the development of urgently required treatment options.

## Material and Methods

### Ethics statement

This exploratory study is in compliance with the Helsinki Declaration (Ethical Principles for Medical Research Involving Human Subjects) and was conducted in accordance with the guidelines of research boards at the study site after written informed consent. Recruitment of Long COVID Syndrome patients was approved by the local ethics committee of the Medical University of Vienna under the number EC#1280/2020, while recruitment of the COVID19 recovered and the healthy/vaccinated control groups was approved by the local ethics committee of the Medical University of Vienna under the number EC#2262/2020. The recruitment of an independent cohort including 10 healthy subjects which had not been exposed to SARS-CoV-2 receiving tablets containing 870mg Omega-3 (420 mg EPA and 330 mg DHA) twice daily in a prospective study design for one week was approved by the local ethics committee of the Medical University of Vienna under the number EC#2250/2020.

### Patient recruitment and sample acquisition

All study participants were recruited in the timeframe between May and July 2021. LCS patients were recruited at the outpatient ward of the Division of Cardiology of the Department for Internal Medicine II of the Medical University of Vienna, Austria. The age and gender matched recovered/healthy and the healthy/vaccinated study groups were recruited among volunteers after calls at the Vienna General Hospital/Medical University of Vienna and the University of Applied Sciences, Vienna, Austria. All vaccinated participants had received two doses of either the vector-based vaccine Vaxzevria (Astra Zeneca, Oxford, UK) or the mRNA-based vaccine Comirnaty (Pfizer, New York City, NY, USA). Serum and EDTA-anticoagulated plasma were obtained by peripheral venous blood draw. Serum samples were left for 15 min to allow for clotting before centrifugation for 15 min at 1500 g at 4 °C while EDTA plasma samples were immediately centrifuged after blood collection. Following these steps, all samples were immediately frozen at -80 °C until analyses.

Regarding the fourth cohort of 10 healthy subjects which had not been exposed to SARS-CoV-2, tablets containing 870mg Omega-3 (420 mg EPA and 330 mg DHA) were administered twice daily in a prospective study design for one week. EDTA-anticoagulated plasma was obtained before start of intake and after intake of the last dose. EDTA plasma samples were immediately centrifuged after blood collection and frozen at -80 °C until analyses.

### Determination of anti SARS-CoV-2 antibody status

Antibody levels against SARS-CoV-2 Spike protein (anti-S) and Nucleocapsid Protein (anti-N) were determined from sera of the study participants using the Elecsys Anti-SARS-CoV-2 S immunoassay as well as the qualitative Elecsys Anti-SARS-CoV-2 immunoassay (detecting SARS-CoV-2 N protein) on a cobas e801 analyzer (Roche Diagnostics, Rotkreuz, Switzerland). Analyses were performed at the diagnostic laboratory at the Department of Laboratory Medicine, Medical University of Vienna (certified acc. to ISO 9001:2015 and accredited acc. to ISO 15189:2012).

### Determination of routine laboratory parameters

Total protein, albumin, ferritin, CRP, HDL, LDL, LP(a) were determined using standard routine diagnostic tests on a cobas e801 analyzer (Roche) at the diagnostic laboratory at the Department of Laboratory Medicine, Medical University of Vienna.

### Determination of serum cytokines

Undiluted blood serum samples were analyzed using the ProcartaPlex™ Multiplex Immunoassay (Human immune monitoring 65 Plex, Thermo Fisher Scientific, Reference Number MAN0017980) according to manufacturer’s instruction. Of the 65 analytes, 51 were below the lower limit of quantification in >95% of all samples (irrespective of group) and were therefore excluded from further analysis. Measurements and analysis of all Human ProcartaPlex Immunoassays were performed on a Luminex 200 instrument (Luminex Corp., Austin, Tx, USA) as described in detail before (*2*).

### Plasma proteomics

For untargeted plasma proteomics analyses, EDTA-anticoagulated plasma samples were diluted 1:20 in lysis buffer (8 M urea, 50 mM TEAB, 5 % SDS), heated at 95 °C for 5 min prior the protein concentration was determined using a BCA assay. For enzymatic protein digestion, 20 µg of protein was used and the ProtiFi S-trap technology (*54*) applied. Briefly, solubilized protein was reduced and carbamidomethylated by adding 64 mM dithiothreitol (DTT) and 48 mM iodoacetamide (IAA), respectively. Prior to sample loading onto the S-trap mini cartridges, trapping buffer (90 % v/v methanol, 0.1M triethylammonium bicarbonate) was added. Thereafter, samples were thoroughly washed and subsequently digested using Trypsin/Lys-C Mix at 37 °C for two hours. Finally, peptides were eluted, dried and stored at -20 °C until LC-MS analyses.

LC-MS/MS analysis was performed as described previously (*55*). In short, Reconstitution of dried peptide samples was achieved by adding 5 µL of 30 % formic acid (FA) containing 4 synthetic standard peptides and subsequent dilution with 40 µL of loading solvent (97.9 % H2O, 2 % ACN, 0.05 % trifluoroacetic acid). Thereof, 1 µL were injected into the Dionex Ultimate 3000 nano high performance liquid chromatography (HPLC)-system (Thermo Fisher Scientific). In order to pre-concentrate peptides prior to chromatographic separation, a pre-column (2 cm□×□75 µm C18 Pepmap100; Thermo Fisher Scientific) run at a flow rate of 10 µL/min using mobile phase A (99.9 % H2O, 0.1 % FA) was used. The subsequent peptide separation was achieved on an analytical column (25 cm □ x □ 75 µm 25 cm Aurora Series emitter column (Ionopticks)) by applying a flow rate of 300 nL/min and using a gradient of 7 % to 40 % mobile phase B (79.9 % ACN, 20 % H2O, 0.1 % FA) over 43 min, resulting in a total LC run time of 85 min including washing and equilibration steps. Mass spectrometric analyses were performed using the timsTOF Pro mass spectrometer (Bruker) equipped with a captive spray ion source run at 1650 V. Further, the timsTOF Pro mass spectrometer was operated in the Parallel Accumulation-Serial Fragmentation (PASEF) mode and a moderate MS data reduction was applied. A scan range (m/z) from 100 to 1700 to record MS and MS/MS spectra and a 1/k0 scan range from 0.60 – 1.60 V.s/cm2 resulting in a ramp time of 100 ms to achieve trapped ion mobility separation were set as further parameters. All experiments were performed with 10 PASEF MS/MS scans per cycle leading to a total cycle time of 1.16 s. Furthermore, the collision energy was ramped as a function of increasing ion mobility from 20 to 59 eV and the quadrupole isolation width was set to 2 Th for m/z < 700 and 3 Th for m/z > 700.

Subsequent LC-MS data analysis was performed using the publicly available software package MaxQuant 1.6.17.0 running the Andromeda search engine (*56*). Protein identification as well as label-free quantification (LFQ) was achieved by searching the raw data against the SwissProt database “homo sapiens” (version 141219 with 20380 entries). General search parameter included an allowed peptide tolerance of 20 ppm, a maximum of 2 missed cleavages, carbamidomethylation on cysteins as fixed modification as well as methionine oxidation and N-terminal protein acetylation as variable modification. A minimum of one unique peptide per protein was set as search criterium for positive identifications. In addition, the “match between runs” option was applied, using a 0.7 min match time window and a match ion mobility window of 0.05 as well as a 20 min alignment time window and an alignment ion mobility of 1. An FDR≤0.01 was set for all peptide and protein identification.

LC-MS data evaluation as well as statistical analysis was accomplished using the Perseus software (version 1.6.14.0) (*57*). First, identified proteins were filtered for reversed sequences as well as common contaminants and annotated according to the different study groups. Prior to statistical analysis, LFQ intensity values were transformed (log2(x)), and proteins were additionally filtered for their number of independent identifications (a minimum of 5 identifications in at least one group). Afterwards, missing values were replaced from a normal distribution (width: 0,3; down shift: 1,8). Two-sided t-tests as well as statistics for volcano plots were performed applying an FDR of 0.05 and a S0 of 0.1, whereby S0 controls the relative importance of t-test p-value and difference between the means. Histograms were generated using GraphPad Prism Version 6.07 (2015).

### Plasma lipidomics

The analysis of fatty acids and oxylipins was performed essentially as described (*58*). Frozen EDTA-anticoagulated plasma was freshly thawed on ice. For precipitation of proteins, plasma (300 µL) was mixed with cold EtOH (1.2 mL, abs. 99%, -20°C; AustroAlco) including an internal standard mixture of 12S-HETE-d8, 15S-HETE-d8, 5-Oxo-ETE-d7, 11,12-DiHETrE-d11, PGE2-d4 and 20-HETE-d6 (each 100 nM; Cayman Europe, Tallinn, Estonia). The samples were stored over-night at -20°C. After centrifugation (30 min, 5000 rpm, 4°C), the supernatant was transferred into a new 15 mL Falcon™ tube. EtOH was evaporated *via* vacuum centrifugation at 37°C until the original sample volume (300 µl) was restored. For solid phase extraction (SPE) samples were loaded onto preconditioned StrataX SPE columns (30 mg mL−1; Phenomenex, Torrance, CA, USA) using Pasteur pipettes. After sample loading, the SPE columns were washed with 5 mL of MS grade water and eluted with ice-cold MeOH (500 µL; MeOH abs.; VWR International, Vienna, Austria) containing 2% formic acid (FA; Sigma-Aldrich). MeOH was evaporated using a gentle nitrogen stream at room temperature and the dried samples were reconstituted in 150 µL reconstitution buffer (H_2_O:ACN:MeOH + 0.2% FA–vol% 65:31.5:3.5) containing an internal standard mixture of 5S-HETE-d8, 14,15-DiHETrE-d11 and 8-iso-PGF2a-d4 (10–100 nM; Cayman Europe, Tallinn, Estonia). The samples were then transferred into an autosampler held at stored at 4°C and subsequently measured via LC-MS/MS.

For LC-MS analyses, analytes were separated using a Thermo Scientific™ Vanquish™ (UHPLC) system equipped with a Kinetex® C18-column (2.6 μm C18 100 Å, LC Column 150 × 2.1 mm; Phenomenex®) applying a gradient flow profile (mobile phase A: H_2_O + 0.2% FA, mobile phase B: ACN:MeOH (vol% 90:10) + 0.2% FA) starting at 35% B and increasing to 90% B (1–10 min), further increasing to 99% B within 0.5 min and held for 5 min. Solvent B was then decreased to the initial level of 35% within 0.5 min and the column was equilibrated for 4 min, resulting in a total run time of 20 min. The flow rate was kept at 200 μL min−1 and the column oven temperature at 40°C. The injection volume was 20 µL and all samples were analysed in technical duplicates. The Vanquish UHPLC system was coupled to a Q Exactive™ HF Quadrupole-Orbitrap™ high-resolution mass spectrometer (Thermo Fisher Scientific, Austria), equipped with a HESI source for negative ionization to perform the mass spectrometric analysis. The MS scan range was 250-700 m/z with a resolution of 60,000 (at m/z 200) on the MS1 level. A Top 2 method was applied for fragmentation (HCD 24 normalized collision energy), preferable 33 m/z values specific for well-known eicosanoids and precursor molecules from an inclusion list. The resulting fragments were analysed on the MS2 level at a resolution of 15,000 (at m/z 200). Operating in negative ionization mode, a spray voltage of 2.2 kV and a capillary temperature of 253 °C were applied. Sheath gas was set to 46 and the auxiliary gas to 10 (arbitrary units).

For subsequent data analysis, raw files generated by the Q Exactive™ HF Quadrupole-Orbitrap™ high-resolution mass spectrometer were checked manually via Thermo Xcalibur™ 4.1.31.9 (Qual browser) and compared with reference spectra from the Lipid Maps depository library from July 2018 (*59*). Peak integration was performed using the TraceFinder™ software package (version 4.1—Thermo Scientific, Vienna, Austria). Principal Component Analysis and volcano plots were generated using the Perseus software (version 1.6.14.0) applying an FDR of 0.05 (*57*). Therefore, data were normalised to the internal standards and missing values were replaced by a constant which corresponds to the half of the lowest normalized area under the curve (nAUC) value of each individual eicosanoid. Histograms were generated using GraphPad Prism Version 6.07 (2015). The ratio of Ω-3/Ω6 fatty acids depicted in Figure 2 reflects the ratio of the omega-3 polyunsaturated fatty acids eicosapentaenoic acid (EPA) and docosahexaenoic (DHA) to the omega-6 polyunsaturated fatty acids arachidonic acid (AA) and dihomo-gamma-linolenic acid (DGLA).

### Plasma metabolomics

EDTA plasma samples (10 µL) of study participants were analyzed by a targeted metabolomic assay. Targeted metabolomics experiments were conducted using the MxP® Quant 500 Kit (Biocrates Life Sciences AG, Innsbruck, Austria). We detected 630 analytes, including 40 acylcarnithines, 1 alkaloid, 1 amine oxide, 50 amino acid related metabolite, 14 bile acids, 9 biogenic amines, 7 carboxylic acids, 28 ceramides, 22 cholesteryl esters, 1 cresol, 44 diacylglycerols, 8 dihydroceramides, 12 fatty acids, 90 glycerophospholipids, 34 glycosylceramides, 4 hormones, 4 indole derivatives, 2 nucleobase related metabolites, 15 sphingolipids, 242 triacylglycerols, the sum of hexoses and 1 vitamin/cofactor. A total of 474 metabolites showed signal intensities within the quantification window and were further evaluated. Measurements were carried out using LC-MS and flow injection (FIA)-MS analyses on a Sciex 6500+ series mass spectrometer coupled to an ExionLC AD chromatography system (AB Sciex, Framingham, MA, USA), utilizing the Analyst 1.7.1 software with hotfix 1 (also AB SCIEX). All required standards, quality controls and eluents were included in the kit, as well as the chromatographic column for the LC-MS/MS analysis part. Phenyl isothiocyanate (Sigma-Aldrich, St. Louis, USA) was purchased separately and was used for derivatization of amino acids and biogenic amines according to the kit manual. Preparation of the measurement worklist as well as data validation and evaluation were performed with the software supplied with the kit (MetIDQ-Oxygen-DB110-3005, Biocrates Life Sciences). Principal Component Analysis and volcano plots were generated using the Perseus software (version 1.6.14.0) applying an FDR of 0.05 (*57*). Histograms were generated using GraphPad Prism Version 6.07 (2015). The heatmap including 126 triacylglycerols (Supplementary Table S4) was generated by dividing the concentration of each lipid through the average concentration of this lipid over all samples using Excel.

### Cell culture and differentiation of U937 cells

U937 cell line was cultured in RPMI medium (1X with L-Glutamine; Gibco, Thermo Fischer Scientific, Austria) supplemented with 1% Penicillin/Streptomycin (Sigma-Aldrich, Austria) and 10% Fetal Calf Serum (FCS, Sigma-Aldrich, Austria) in T25 polystyrene cell culture flasks for suspension cells (Sarstedt, Austria) at 37 °C and 5% CO_2_. Cells were counted using a MOXI Z Mini Automated Cell Counter (ORFLO Technologies, USA) using Moxi Z Type M Cassettes (ORFLO Technologies, USA). Based on this, 2 × 10^6^ cells were used for each differentiation approach and seeded in T25 polystyrene cell culture flasks with cell growth surface for adherent cells (Sarstedt, Austria).

Differentiation of U937 cells into M1-like macrophages was induced by adding 100 ng/mL Phorbol 12-myristate 13-acetate (PMA, ≥99%, Sigma-Aldrich, Austria) (d0). After 48h (d2), the cell culture medium was exchanged and fresh full medium supplemented with 100 ng/mL LPS (Lipopolysaccharides from Escherichia coli 055:B5, γ-irradiated, BioXtra, Sigma-Aldrich, Austria) was added. Again, after 48h (d4), M1-like macrophages were washed twice with PBS and further incubated with 3 mL of serum free RPMI for 4h. Thereafter, supernatants were harvested, precipitated using 12 mL cold EtOH (abs. 99%, -20°C; AustroAlco) including an internal standard mixture of 12S-HETE-d8, 15S-HETE-d8, 5-Oxo-ETE-d7, 11,12-DiHETrE-d11, PGE2-d4 and 20-HETE-d6 (each 100 nM; Cayman Europe, Tallinn, Estonia) and stored at -20 °C until further processing. M1-like macrophages were lysed in 200 µL of a 4% SDC buffer containing 100 mM Tris-HCl (pH 8.5), immediately heat-treated at 95°C for 5 minutes, ultra-sonicated and stored at - 20 °C until further processing.

M2-like macrophage differentiation of U937 cells was achieved by first adding 100 ng/mL PMA (d0) for 24 h to the full medium. Afterwards, 50 ng/mL M-CSF (ImmunoTools, Friesoythe, Germany) were directly added to the culture medium (d1) for a total of 72 h before the medium was exchanged and cells cultivated in fresh full medium supplemented again with 50 ng/mL M-CSF (d4) for another 72 h. After this (d6), cells were incubated in fresh medium containing 20 ng/mL IL-4 (ImmunoTools) for 24h to induce the M2-like phenotype. At day 7 (d7) of the differentiation process, M2-like macrophages were washed twice with PBS and further incubated with 3 mL of serum free RPMI for 4h. Sample harvesting was performed as described above. M1-like macrophage differentiation as well as M2-like macrophage differentiation of U937 cells were carried out in triplicates.

### Sample preparation and LC-MS analyses of M1-like and M2-like macrophages

Precipitated FCS-free cell supernatants were centrifuged (30 min, 5000 rpm, 4°C) and the supernatant was transferred into new 15 mL Falcon™ tubes for lipid extraction as described above. The protein pellet corresponding to the secreted proteins was dissolved in 4% SDC buffer containing 100 mM Tris-HCl (pH 8.5), ultra-sonicated and heat-treated at 95°C for 5 minutes. Protein concentration of supernatants as well as cell lysates was determined using a BCA assay. For proteomic analyses, an adapted version of the EasyPhos workflow was applied (*60*). In short, 20 µg of protein was reduced and alkylated in one step using 100 mM TCEP and 400 mM 2-CAM, respectively. Subsequent enzymatic digestion was performed with a Trypsin/Lys-C mixture (1:100 Enzyme to Substrate ratio) at 37 °C for 18 h. For desalting, peptide solution was first dried to approximately 20 µL, mixed with loading buffer containing 1 % TFA in isopropanol and loaded on SDB-RPS StageTips. After washing twice, peptides were eluted with 60 % ACN and 0.005 % ammonium hydroxide solution, dried and stored at -20 °C until LC-MS analyses.

LC-MS analyses of the supernatants of M1-like and M2-like macrophages were carried out as describe above (plasma proteomics and plasma lipidomics) with slight adaptions regarding the proteomics analyses. In case of secretome analysis 5 µL of resuspended sample were injected into the Dionex Ultimate 3000 nano high performance liquid chromatography (HPLC)-system (Thermo Fisher Scientific) but using the same gradient as for plasma proteomics. Regarding the proteomic analyses of cell lysates, again 5 µL of resuspended sample were injected but an adapted LC-gradient from 8 % to 40 % mobile phase B over 90 min, resulting in a total LC run time of 135 min including washing and equilibration steps, was applied.

## Data Availability

All data produced in the present study are available upon reasonable request to the authors.

## List of Supplementary Materials

**Supplementary Figure S1**: Volcano plots displaying significant differences (marked in red) in

**(A)** proteins, **(B)** eicosanoids and **(C)** metabolites between the different study groups (long-covid patients (long-covid), fully recovered patients (recovered), vaccinated healthy controls (healthy))

**Supplementary Figure S2**: Lactate concentration in plasma of healthy controls (H), recovered (R) and long-covid (LC) patients.

**Supplementary Table S1**: Identified proteins including statistics

**Supplementary Table S2**: Identified eicosanoids including statistics

**Supplementary Table S3**: Identified metabolites including statistics

**Supplementary Table S4**: Heat map data matrix

## Acknowledgments

This work was supported by the University of Vienna and by the Joint Metabolome Facility (University of Vienna, Medical University of Vienna), member of the VLSI (Vienna Life Science Instruments).

## Author contributions

Conceptualization: JK, MG, KS, CG

Methodology: KS, CG

Investigation: AB, GH, SMM, TF, AK, LS, BR, DS, GG

Visualization: AB, GH, SMM

Funding acquisition: JK, KS

Patient recruitment and clinical assessment: MH, TS, AS, EH, MG

Supervision: AB, CG

Writing – original draft: CG

Writing – review & editing: JK, AB, KS, CG

## Data and materials availability

All data produced in the present study are available upon request to the authors.

