## Supplementary figures and images for "Multi-omics provide evidence for an anti-inflammatory immune signature and metabolic alterations in patients with Long COVID Syndrome – an exploratory study"

### Supplementary Figure S1

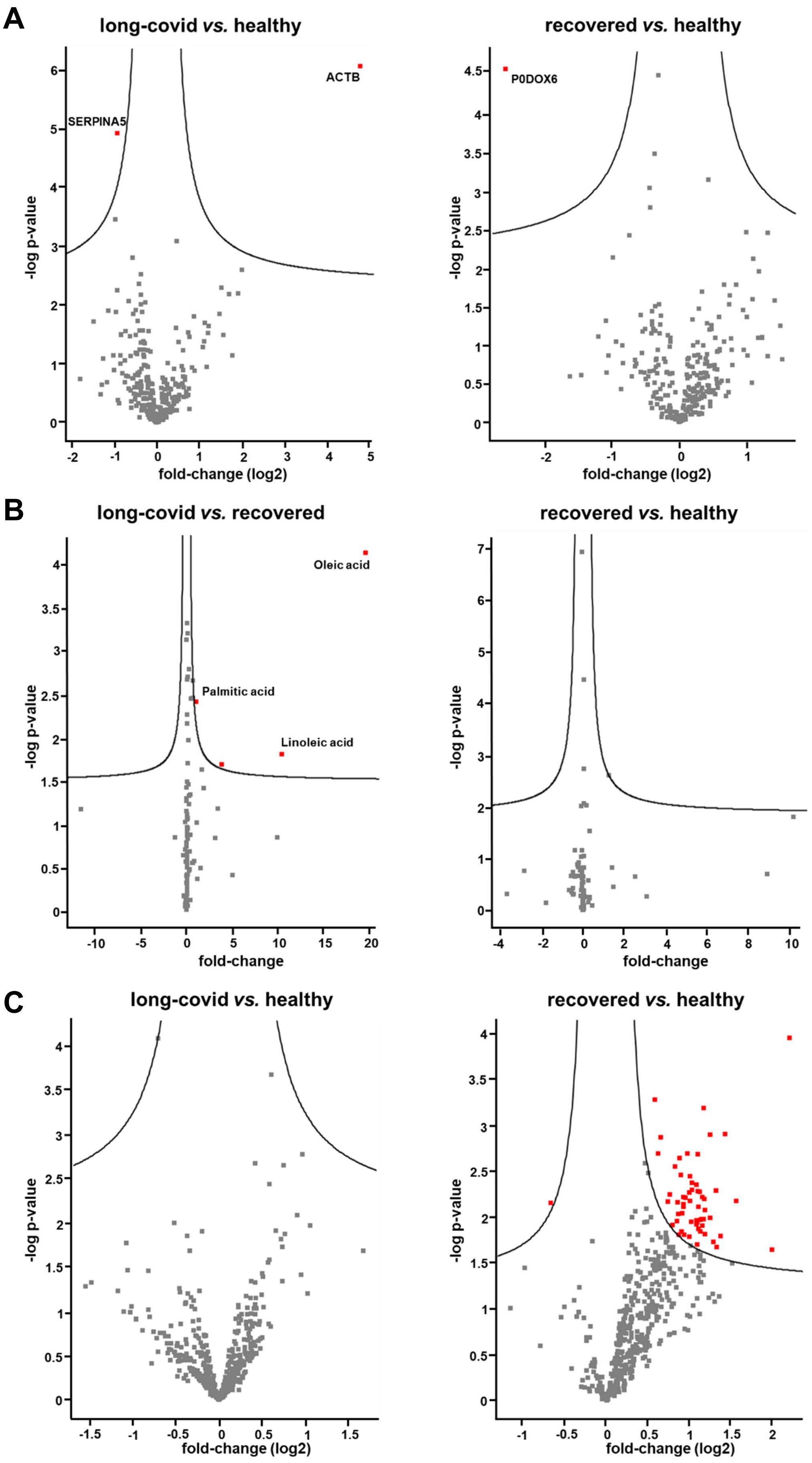


**Supplementary Figure S1**

### Supplementary Figure S2

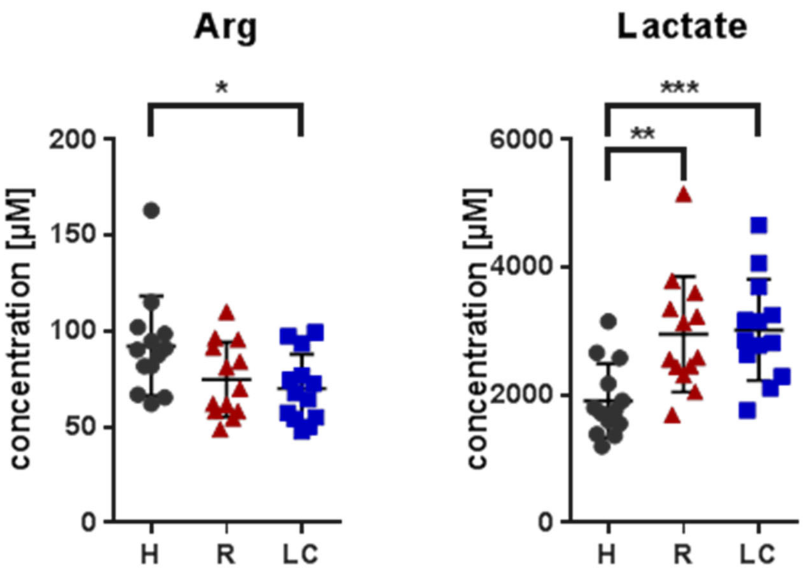


**Supplementary Figure S2**
